# Rehabilitation in Survivors of COVID-19 (RE2SCUE): a nonrandomized, controlled, and open protocol

**DOI:** 10.1101/2021.09.06.21262986

**Authors:** Maria Cristine Campos, Tatyana Nery, Ana Cristina de Bem Alves, Ana Elisa Speck, Danielle Soares Rocha Vieira, Ione Jayce Ceola Schneider, Maria Paula Pereira Matos, Livia Arcêncio, Aderbal Silva Aguiar

**Affiliations:** Exercise Biology Lab (Labioex), Department of Health Sciences, Federal University of Santa Catarina, Araranguá, Brazil; Cardiovascular and Respiratory Assessment and Rehabilitation Lab (LaCOR), Department of Health Sciences, Federal University of Santa Catarina, Araranguá, Brazil; Epidemiological Research Lab (LabEpi), Department of Health Sciences, Federal University of Santa Catarina, Araranguá, Brazil; University of South Santa Catarina (Unisul), Psychology College, Santa Catarina, Tubarão, Brazil

**Keywords:** Rehabilitation, Coronavirus, COVID-19, Respiratory Tract Diseases, Exercise therapy

## Abstract

**Objective:** This study aimed to evaluate the effects of physical rehabilitation for adults with sequelae after COVID-19.

**Methods:** This clinical, nonrandomized, controlled, and open study will examine 82 participants who have met the inclusion criteria and who will be divided into treatment and control groups according to participant preference. The intervention group will receive face-to-face care; the control group will receive remote educational guidance for 8 weeks, with pre-post evaluations. The primary outcomes are dyspnea, fatigue, and exercise capacity; the secondary outcomes are lung function, heart rate variability, handgrip strength, knee extensor strength and electrical activity, physical activity, functional limitation, cognitive function, depression and anxiety, and biochemical measures of hypoxia, inflammation, oxidative stress, blood glucose, and lactate blood tests. The survey will follow the *Standard Protocol Items for Randomized Trials* guidelines, and the results will be reported according to the *Consolidated Standards of Reporting Trials* guidelines. Effects will be assessed based on the intent-to-treat data collected. Analysis of covariance will be used for the initial and final evaluations, with a significance level of 5%.

**Results and Conclusions:** The results will show the effectiveness of rehabilitation in adults with post-COVID-19 sequelae.

**Impact:** Fatigue, dyspnea, cough, and muscle and joint pain are common sequelae of post-COVID-19 syndrome. Physical rehabilitation is one modality for treating these sequelae. This protocol can provide a treatment model for patients with post-COVID-19 sequelae.

## Introduction

Coronavirus disease 2019 (COVID-19) is a global public health emergency^(1)^ caused by coronavirus 2 that affects multiple organs and systems^(2)(3)(4)(5)(6).^ Similar to other infectious viral conditions, COVID-19 has caused persistent sequelae^(5)^ regardless of disease severity^(7)(8).^ Post-acute symptoms include fatigue, headache, attention disorders, hair loss, dyspnea, cough, ageusia, anosmia, muscle and joint pain, and chest pain, persistent in 80% of individuals after the disease^(7)(9)^.

A prospective cohort study with 1,733 participants has shown that fatigue was the most reported symptom (63%), followed by difficulty sleeping, anxiety, and depression^(10)^. Furthermore, in nonhospitalized patients, dyspnea, fatigue, anosmia, and ageusia may persist for 4−7 months post-infection^(8)^. The late effects and presence of long-lasting symptoms characterize a post-acute COVID-19 syndrome^(5)(9)(11)^.

Impaired health and reduced functional capacity have demonstrated COVID-19’s impact and the need to develop a safe, efficient rehabilitation protocol to recover function and social, professional, and family life^(12)^. Rehabilitation after recovery from acute COVID-19 has been recommended^(1)(13)(14)(15)(16)(17)^ for different degrees of respiratory, physical, and psychological dysfunctions^(1)(18)^. The main rehabilitation objectives are to reduce dyspnea, improve functional capacity, reduce anxiety and depression, treat complications resulting from immobility, and facilitate return to social and work activities^(1)(13)^.

Some studies have shown that physical exercise can improve exercise capacity, reduce fatigue and respiratory symptoms, and improve cognition^(19)^, quality of life, and anxiety in these patients^(20)^. However, there is little evidence for rehabilitation safety and effectiveness in post-COVID-19 syndrome. Therefore, this study will investigate the effects of a rehabilitation protocol for adults and older people with post-acute COVID-19 syndrome who are in an extra-hospital environment that is centered on physical exercise, patient education, and lifestyle changes.

## Methods

### Study design

This study is clinical, nonrandomized, controlled, and open. The research will follow the *Standard Protocol Items for Randomized Trials* (SPIRIT)^(21)^, and the results will be reported according to the *Consolidated Standards of Reporting Trials*^(22)^.

### Participant allocation

A researcher will manage the participant allocation. The selected volunteers will be allowed to choose to participate in either the control (remote educational guidance) or intervention group (face-to-face physical rehabilitation) according to their preferences and availabilities. This nonrandomized allocation was determined after the pilot study indicated low adherence by the control group.

### Sample size calculation

The sample size calculation is based on the functional capacity results assessed by the 6-min walking test from the 2020 study by Kai Liu et al.^(20)^. Considering a power of 80%, a significance level of 5%, and a 10% loss to follow-up, the estimation was 41 participants per group.

### Participant recruitment

The clinical trial will be conducted at the Hospital Regional de Araranguá (HRA), a regional hub in the southern state of Santa Catarina, Brazil. Recruitment will be carried out from the COVID-19 register of the region’s Health Department through phone and press calls and social networks. Participant inclusion and exclusion criteria are shown in Table 1.

**Table 1.** Study participants’ inclusion and exclusion criteria. Legend: RT-PCR, reverse-transcriptase polymerase chain reaction; CVD, cardiovascular disease; COPD, chronic obstructive pulmonary disease; NYHA, New York Heart Association.

| STUDY PERIOD |  |  |  |  |
| --- | --- | --- | --- | --- |
|  | Enrolment | Allocation | Post-allocation | Close-out |
| TIMEPOINT | $-t_1$ | 0 | $t_1$ | $t_8$ |
| ENROLMENT: |  |  |  |  |
| Eligibility screen | X |  |  |  |
| Informed consent |  | X |  |  |
| Allocation |  | X |  |  |
| INTERVENTIONS: |  |  |  |  |
| *[Intervention]* |  |  | ↔ |  |
| *[Control]* |  |  | ↔ |  |
| ASSESSMENTS: |  |  |  |  |
| *[Primary and Secondary outcomes]* |  |  | X | X |

### Participant discontinuity and adherence strategies

Participants will be discontinued from treatment if a new adverse event, such as an additional COVID-19 infection, significant hemodynamic changes, and/or musculoskeletal injury prevent physical training.

A follow-up every 15 days will reinforce healthy habits and guidance on physical activities for the control group. At treatment’s end, the participants will be invited to contribute an individualized satisfaction survey about their intervention. Justifications for study withdrawals will be reported. Participants can choose not to participate in some measures as registered on an informed-consent form.

### Group evaluation and assessment

Trained physiotherapists will evaluate all participants pre- and posttreatment. Participants will receive care for 8 consecutive weeks (Fig. 1). Moreover, participants will undergo an electrocardiogram examination before treatment, with results reported by a cardiologist.

**Figure 1.** Schedule of participant enrollment, interventions, and assessments. Legend: −t_1_, fulfillment of the inclusion criteria; t_1_, initial evaluation; t_2_, evaluation after 8 weeks of treatment; t_8_, final evaluation.

The intervention group (n = 41) will undergo 8 consecutive weeks of physical rehabilitation, 2×/week, with an average duration of 60 min/session. The control group (n = 41) will receive educational guidance and assistance via video call, telephone, or text message and will also be instructed to improve their physical activity (PA).

### Intervention group treatment

Participants in the intervention group will undergo rehabilitation supervised by physiotherapists. Hemodynamic parameters (blood pressure [BP], heart rate [HR]), peripheral oxygen saturation, and perceived exertion will be continuously monitored.

Aerobic exercise will be performed on a treadmill at moderate intensity at 60%−75% of the reserve HR calculated using the Karvonen method and 4−6 on a modified Borg scale rating of perceived exertion^(23)^. The initial treadmill speed will be 75% of the average speed reached in the incremental shuttle walk test (ISWT)^(24)^. Participants will perform 5 min of warming up and cooling down and 30 min of aerobic exercise.

The resistance exercises will be applied in 3 × 10 sets, with an initial 80% load of ten maximum repetitions for trunk and back and upper and lower limbs, with 1–2-min intervals between sets. Finally, the exercised muscles will be slightly stretched (30 s) at the end of the session. Figure 2 illustrates the intervention group’s exercise protocol.

**Figure 2.** Illustration of the intervention group’s exercise protocol. The images illustrate (a) aerobic training on a treadmill, (b−f) active stretching, and (g−l) resistance exercises for the arms, legs, and chest muscles. *Source:* Authors (2021)

If necessary, breathing exercises for expansion improvement and bronchial hygiene will be performed.

### Control group treatment

A booklet with general guidelines on breathing etiquette, nutritional information, water intake, energy conservation techniques, breathing exercises, and PA will be provided to participants in the control group. Every fortnight, participants will receive a video call to reinforce the guidelines and monitor health status. For more details on the guidelines and SPIRIT Checklist, see the supplementary materials 1 and 2, respectively.

### Outcome measures

#### Primary measures

The primary outcomes of the study are dyspnea, fatigue, and exercise capacity. Dyspnea symptoms will be assessed by the Medical Research Council scale. The Modified Pulmonary Functional Status Dyspnea Questionnaire will be used to evaluate dyspnea and fatigue in activities of daily living (ADLs)^(25)^.

Exercise capacity will be assessed using the ISWT, a progressive walking test with sound cues (beeps) and controlled speed, which is a valid and reliable evaluation of the functional capacity of respiratory patients^(26)^. The primary outcome will be the distance covered^(26)^. Two measurements will be taken, with the longest distance considered due to the learning effect (26)(27).

#### Secondary measures

Secondary outcome measures will be lung function, handgrip strength, knee extensor strength and electrical activity, functional status, anxiety and depression, PA, HR variability (HRV), cognitive function, and hypoxia, inflammation, oxidative stress, and blood lactate and glucose. A more detailed description of the measurements for each outcome is provided below. Figure 3 presents the model for collecting and analyzing the study data and the research development.

**Figure 3.**
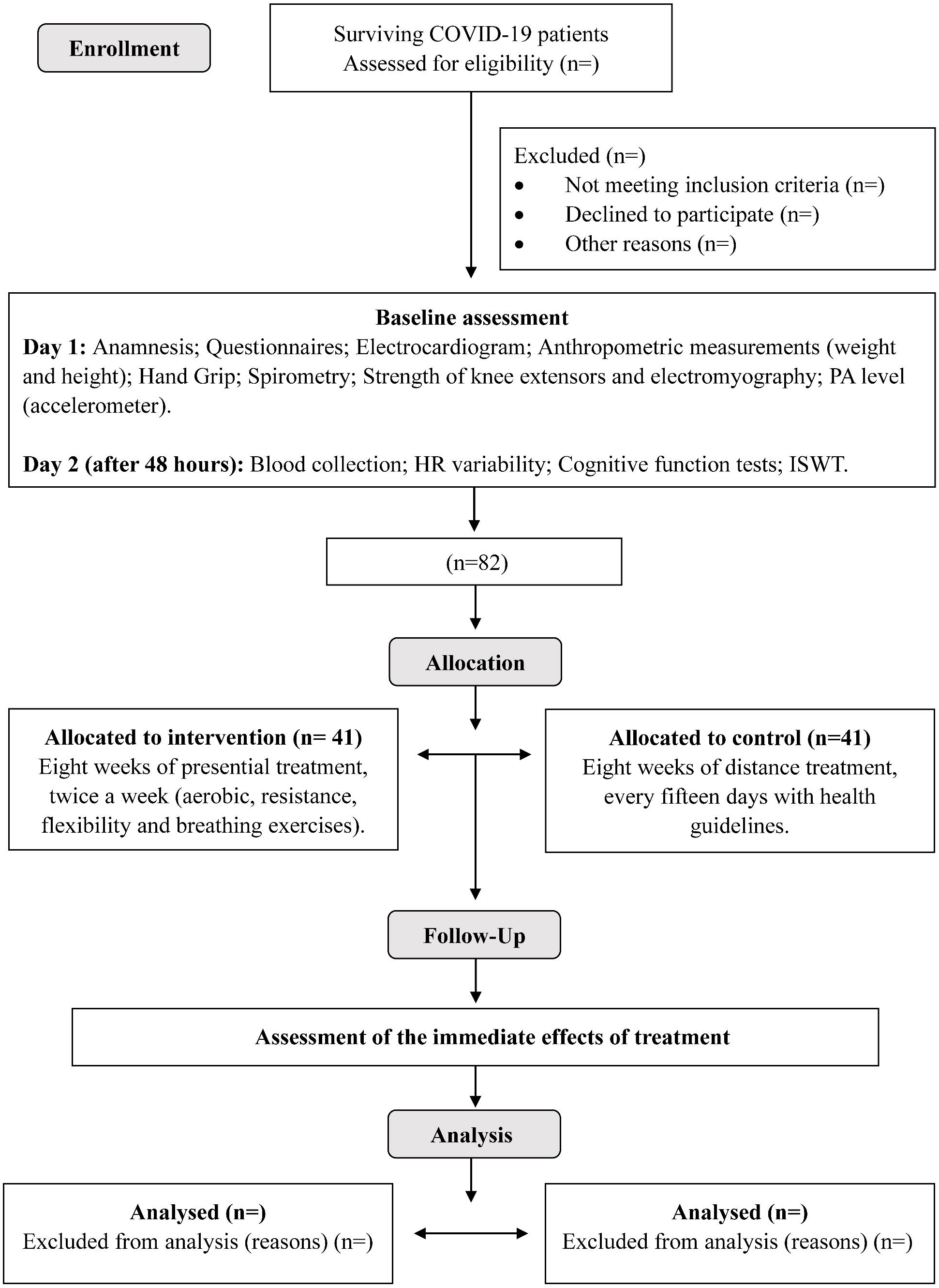
Flowchart of the study conduction process. *Source*: Authors (2021).

##### Lung function

Spirometry will evaluate lung function following the Brazilian and international guidelines^(28)(29)^. The following variables will be considered: forced vital capacity (FVC), forced expiratory volume in 1 s, peak expiratory flow, and the average forced expiratory flow between 25% and 75% of the FVC (25%−75%)^(30)^. The 2007 predicted values by Pereira et al. will be used^(31)^.

##### Handgrip strength

Following the American Society of Hand Therapy recommendations, handgrip strength will be assessed using a Jamar® dynamometer^(32)^. The participants will be seated in a chair with a backrest and without upper-limb support, feet flat on the floor, dominant shoulder in the adducted position, elbow flexed at 90°, and forearm in neutral^(33)^. Three maximum isometric contractions held for 6 s with an interval of 1 min will be collected.

##### Knee extensor strength and electrical activity

Knee extensor strength associated with surface electromyography will be evaluated through the maximum voluntary isometric contraction (MVIC) of the dominant lower limb^(34)(35)^. The NewMiotol digital converter will register the traction force in the Miograph software (force load cell model; Miotec Biomedical Equipment, Porto Alegre, Brazil). The electrodes will be attached to the quadriceps according to the Surface ElectroMyoGraphy for the Non-Invasive Assessment of Muscles specifications.

Participants will perform the test sitting in a chair with a backrest, 90° of hip flexion, 60° of knee flexion, and arms to the chest. With instruction and verbal encouragement, participants will perform as much isometric strength as possible for 6 s^(36)(37)^. The muscle fatigue test will be considered 50% of the MVIC, with ankle weights. Participants will perform the test until fatigue, defined by the loss of tibial tuberosity contact with a metal rod. The isometric contraction time until the point of fatigue will be registered^(38)^.

##### PA level

PA level will be evaluated by the ActiGraph® wGT3X-BT accelerometer (Pensacola, FL, USA). Participants will be instructed to use the instrument for 7 consecutive days. Data will be collected considering a sampling rate of 30 Hz and analyzed using a 60-s epoch. Periods of nonuse, defined as 60 min of consecutive 0 counts, with a peak acceleration tolerance of 2 min, will be excluded^(39)^. The PA level will be classified based on the adult recommendations^(40)^.

The data validated will be for those with 3 days of measurements, including 1 weekend day, with at least 10 h of daily use (or 600 min)^(41)^. Participants will receive written instructions, along with a diary for recording moments of nonuse.

##### Functional status

Functional status will be assessed by the Post-COVID-19 Functional Status Scale, composed of five levels (0−4), in which 0 corresponds to *the absence of functional limitation*, and 4 is *severe limitation with the need for assistance in all ADLs*^(42)^.

##### Cognitive function

Cognitive function will be assessed using memory prospective, retrospective, and focused attention tests. The Reminder Prospective Memory Test assesses prospective memory. Participants will be instructed to request the sending of a message by phone (WhatsApp) with the results of their vital signs before starting the field test (ISWT)^(43)^. The Red Pen Prospective Memory Test will instruct the participants to remember to request a red pen to complete the attention test, which is carried out afterward. The red pen will be kept in a drawer, while a blue one will be on the table^(44)(45)^.

The Brazilian version of the Rey Auditory−Verbal Learning Test will be used for retrospective memory assessment. This easy-to-apply test is widely recognized in the neuropsychological literature^(46)^.

Focused attention will be assessed using the Attention Test version R, “d2-R,” which consists of 1 page with 14 lines with the letters *d* and *p* distributed. The target symbol is a *d* with two dashes (d2). The objective is to strike as many targets as possible, constantly moving from left to right of the line, with a time limit of 20 s for each line attempt^(47)^.

##### Anxiety and depression

The assessment of anxiety and depression will be carried out using the Hospital Anxiety and Depression Scale^(48)^. The response scale for each item ranges from 0 to 3, with a total score for each subscale of 21 points^(49)^. Scores of ≥8 indicate anxiety and depression.

Anxiety symptoms will be assessed using the Anxiety Inventory for Respiratory Disease. Each item is scored from 0 (*no anxiety symptoms*) to 3 (*anxiety symptoms almost all the time*) points, with a total score of 0−30. A score of ≥8 suggests the presence of anxiety^(50)^.

##### HRV

HRV^(51)^ will be measured by the V800 HR monitor (Polar Electro OY, Kempele, Finland)^(52)^. Participants will wear a chest strap with an HR sensor and a wristwatch. The HRV recording will be performed with the participants in the supine position for 20 min at rest.

The data will be saved via the Polar Flow web service synchronized with the FlowSync software and analyzed by the Kubios HRV Standard software. Linear methods in time and frequency will be analyzed^(51)^.

##### Biochemical measurements of hypoxia, inflammation, oxidative stress, and blood glucose and plasma lactate

A blood sample will be collected in a specific hospital laboratory. Blood collection will be the first activity of the day, with no prior physical effort or need for fasting.

Subsequently, 10 mL of antecubital venous blood collected in EDTA heparinized tubes will be centrifuged with 6.6 μl of Histopaque® 1119 at 2,000 rpm for 30 min to obtain and store the plasma. Thereafter, leucocytes (next layer) will be transferred to a Falcon tube with 10 mL (qs) of 0.9% saline and centrifuged at 1,500 rpm for 10 min. The leucocyte pellets will then be collected for storage. If the pellet is not available, this process will be repeated with saline 1:10 of H_2_O Milli-Q®.

The last step requires discarding the remaining Histopaque from the primary tubes and adding 10 mL (qs) of 0.9% saline to the blood. After being homogenized 4×, these tubes will be centrifuged at 1,500 rpm for 10 min. After discarding the Histopaque®, the red blood cells will be collected and stored at −80°C, along with the plasma and leucocytes, until biochemical analysis.

Hypoxia (hypoxia factors 1 and 2, shock protein 70) and inflammation (interleukins 1β, 4, 6, and 10) markers will be analyzed with commercial enzyme-linked immunosorbent assay kits.

Capillary blood will be collected at the end of the ISWT to measure blood glucose with the G-TECH Lite monitor and a lactate analyzer (Lactate Detect TD-4261) in mmol/L.

### Data monitoring committee

This study will not have a data monitoring committee, considering that the exercises performed are of low-to-moderate intensity and will be supervised by experts^(16)^. The treatment session will always be held in the hospital’s internal environment, where medical services specialized for urgencies and emergencies are immediately available. In addition, all participants in both groups will be encouraged to report any discomfort during appointments.

### Statistical analysis

A numbered ID will identify participants. Two independent team members will type the data collected on a computer and check for missing or erroneous values. In addition, all paper and electronic files will be kept in a safe place, with access restricted to the research team. The analysis will be carried out by a researcher blinded to the participants’ allocation through the program Stata SE 16.1 (2020; StataCorp. College Station, TX, USA).

A significance level of 5% will be considered. The data will be presented as absolute and relative frequencies and as central tendency and dispersion measures. We will report 95% confidence intervals.

An intention-to-treat analysis will be held^(53)^. An analysis of covariance will be used to compare the control and intervention groups, with adjustments for the variables age, sex, and admission to the intensive care unit. The effect size will be calculated and classified as small (0.20−0.50), average (0.50−0.80), or large (>0.80)^(54)^.

## Discussion

The results will be analyzed, reviewed, and submitted to scientific journals. Semiannual meetings will occur during all study phases to follow up with the Ministry of Health and the CNPq.

This study provides a complete description of the rehabilitation protocol for adult patients with COVID-19 sequelae. The results will demonstrate the effects of rehabilitation, providing assessment and treatment methods and contributing to the public health policies for patients with COVID-19 sequelae and to the clinical decisions of professionals who work directly in their physical rehabilitation.

Considering the impact of post-COVID-19 syndrome on the functionality of the affected patients, especially for fatigue and dyspnea, our study addresses a moderate-intensity exercise treatment as a central component of rehabilitation.

## Data Availability

Data are available through the corresponding author

## Acknowledgments

The authors thank the volunteers, the Hospital Regional de Araranguá (HRA), and the funding sources.

## Ethics Approval

This protocol was approved by the University’s Ethics Research Committee (CAAE: 38682820.0.0000.0121) and is registered with the Brazilian Clinical Trials Registry (U1111-1261-4925). The study will be conducted according to Resolution 466/2012 of the National Health Council. All participants must sign an informed-consent form.

