## Supplementary material for "Rehabilitation in Survivors of COVID-19 (RE2SCUE): a nonrandomized, controlled, and open protocol": Table 1

**Inclusion criteria**

- Patients >18 years old of both sexes diagnosed with COVID-19 in the past 6 months.
- Patients diagnosed with COVID-19 by RT-PCR and/or clinical−epidemiological means.
- Patients having the presence of at least one of the following symptoms: fatigue, dyspnea, muscle pain, joint pain, and cough.
- Patients with CVD receiving regular medical follow-ups.

**Exclusion criteria**

- Patients with respiratory diseases before COVID-19, such as asthma, COPD, or pulmonary fibrosis.
- Patients with moderate-to-severe heart disease (NYHA III or IV).
- Patients with neurological or osteoarticular diseases that prevent study participation.
- Patients who have difficulty understanding the tests performed in the study.
- Patients with cardiovascular symptoms or changes in their electrocardiogram that contraindicates physical exercise (as reported by a cardiologist).
