## supplementary material 2 for "Rehabilitation in Survivors of COVID-19 (RE2SCUE): a nonrandomized, controlled, and open protocol"

Supplementary Material #1

**Post-COVID-19 Guidance Manual**

The guidelines manual was intended for patients in the study's control group. The content consists of essential guidelines for health and self-care, promoting well-being, physical and mental health.

**Respiratory etiquette**

- Personal contact should be avoided (such as hugs and handshakes);
- Whenever you cough or sneeze, cover your mouth and nose with a disposable handkerchief and if not possible, cover it with your forearm;
- Hands should always be sanitized with soap and water or gel alcohol;
- The use of a mask is essential;
- You must be careful to avoid putting your hands on your face;
- A distance of at least two meters must be kept between people.

**Nutritional Guidelines**

- Make fresh or minimally processed foods the basis of your diet.
- Use oils, fats, salt, and sugar in small amounts when preparing meals.
- Limit the use of processed foods by consuming them in small amounts.
- Avoid consuming ultra-processed foods.
- Before placing food in cupboards or pantries, wash containers with soap and water and/or spray 70% alcohol or chlorinated solution.
- Remove fruits, vegetables, and vegetables from packaging and store in fruit bowls. If going to the refrigerator, sanitize first.
- Before consuming raw fruits and vegetables, wash them under running water and sanitize them with sodium hypochlorite 2%. It must always be used diluted, as per the manufacturer's recommendations.

**Water and Hydration**

- During your recovery, proper hydration is essential!
- The amount of water needed per day varies widely and depends on several factors (age, weight, physical activity, etc.)
- Consume at least 2 liters of fluid throughout the day.
- The water ingested must originate from consuming pure water and water contained in food, culinary preparations and is also valid for teas, natural juices, and coconut water.
- Important: If you have any kidney or heart disease that causes restriction of fluid volume, consult your physician.

**Energy conservation**

Energy conservation techniques reduce shortness of breath and fatigue to carry out daily activities. In this section are some examples of these techniques.

- Choose clothes that are comfortable and easy to wear.
- Avoid lifting and carrying heavy objects.
- Take a sit-down shower (plastic chair), especially to wash your hair.
- Sit when brushing your teeth or shaving, and keep your arms supported as much as possible.
- Equipment and utensils must always be within easy reach.

**Breathing exercises**

- Lip Fret Breathing Exercise: Sit in a chair or armchair with your neck and shoulders relaxed. Gently draw air through your nose and also gently release it between your partially closed lips. Mentally count to 2 as you pull in perspective and to 4 as you release air. Necessary: This exercise can be performed during your daily activities and also during physical exercise.
- Diaphragmatic breathing exercise: Sit comfortably in an armchair. Place one hand over your chest and the other hand over your belly. Gently suck in air through your nose as you feel your stomach move outward. Then hold the air in your lungs for 3 seconds and slowly release it between your lips. Repeat 8 to 10 times.

**Physical Exercise**

- Start gradually so your body will adapt.
- Control your breathing during exercise, avoiding holding your breath.
- Drink water during exercise.
- Do not exercise right after a meal.
- Do not exercise if you have a fever, severe headache, severe shortness of breath, palpitations (rapid heart).
- If you do not have physical space at home or equipment, preferably in open, well-ventilated places, and times that have less movement of people, always wear the mask.
- Wear comfortable clothing, sunglasses, and a hat while exercising, and do not exercise on very hot or humid days (northeast wind).
- Follow the intensity scale. The Modified Borg scale serves to assess your feeling of tiredness and shortness of breath. During exercise practice, these sensations should be maintained between the intensity of 2 (mild) to 3 (moderate) for the arm exercises and 3 (middle) for the leg exercises.
- Exercises for the arms. You will do 8 to 10 reps 2 to 3 times. These exercises can be performed 2-3 times a week with at least one rest day in between. For these exercises, your fatigue should be between 2 (mild) and 3 (moderate) on the Borg scale (below). Stop exercising if you experience any abnormal symptoms (chest pain, severe joint pain, dizziness or vertigo, palpitations, headache, severe shortness of breath).
- Standing, position your hands so that their palms face forward; gradually raise your hands towards your chest, and slowly lower them back to the starting point. Pull a little air through your nose before bending your arms and letting it go between your lips throughout the movement.
- Standing, keep your torso straight and slowly raise your arms until they are at shoulder height, with your palms facing the floor. Slowly lower your arms back to your sides. Breathe in through your nose before raising your arms and release it between your lips throughout the movement.
- Sitting, spread your arms wide. You will open and close them. Pull a little air through your nose at first and let it out between your lips as you open and close your arms.
- Exercises to improve your muscle and cardiorespiratory fitness. You will do 8 to 10 reps 2 to 3 times. These exercises can be performed 2-3 times a week with at least one rest day in between. For these exercises, your tiredness should be around 3 (moderate) on the Borg scale. Stop exercising if you experience any abnormal symptoms.
- Sit and stand exercise: Start sitting and then you will rise and set from the chair. Breathe in through your nose and let it out between your lips as you get up and sit back in the chair.
- Calf Exercise: Support yourself on a chair or other surface. Pull a little air through your nose and release it between your lips as you stand on your tiptoes, and return to the starting position.
- Squat exercise: With arms supported on a firm chair or table, perform semi squats and return to starting position. Pull a little air through your nose and release it between your lips as you complete the semi squats.
- Walking exercise: Start with light walking (30 min/day).
- Stationary Gait: Take a walk without leaving your seat—1 to 2-minute sets. Repeat the exercise 5 times.
- Going up and down stairs exercise: In front of a ladder, go up and down the stairs. 1 to 2-minute sets. Repeat the exercise 3 times.

**Stretches**

- Perform these exercises daily, preferably after the workout, to improve your cardiorespiratory fitness. Hold the poses for 30 seconds, with three reps per side. Control your breathing during exercise, exhaling air between your lips.
- Neck Stretch: Sitting, with your spine stretched and supported, place your hand on top of your head, pulling slightly to the right. Return to starting position and repeat on the left side.
- Cross-Arm Stretch: Cross your right arm in front of your body with your elbow straight. Use your left hand to keep it in position. Repeat with left arm straight and right hand holding it in place.
- Leg Stretch: Stand in front of a chair a little less than an arm's length away. Hold the chair with both hands and step forward with your right leg and one step back with your left leg, keeping your feet parallel.
- Leg Stretch: Sitting in a chair, pull your knee toward your torso and hold with your right hand. Then repeat the procedure with your left knee.

**Secretion Removal Techniques**

Techniques to help remove secretions from the lungs if you feel that you have a buildup of secretions.

- Lie on your side and breathe easily three times. Then keep your mouth open and let all the air out slowly through your mouth. Repeat this procedure at least three times. Now lie down on the other side and repeat the process.
- Then sit down, fill your chest with air, and quickly release the air with your mouth open (as if you were going to fog up a mirror). Repeat this procedure 2-3 times, resting between each repetition.
- Next, perform a cough (if the cough is difficult, you can hug a pillow close to your chest to cough harder) and wipe the phlegm off with disposable tissue. If you prefer, you can swallow the phlegm.
- Necessary: Perform this technique 1-2 times a day; wake up and go to bed if you have a buildup of secretions.

**Considerations**

In addition to performing the indicated exercises, it is essential to stay active and reduce sedentary behavior! It is advised that every 30 minutes in a sitting position, perform some activity! Activities that we serve sitting or lying down do not increase the body's energy expenditure, such as watching television, using the computer and cell phone, among others.
